# Vitamin D and the preclinical Alzheimer cognitive composite cognition (PACC) score: a two-sample mendelian randomization study

**DOI:** 10.1101/2022.11.23.22282674

**Authors:** Yann Ilboudo, Satoshi Yoshiji, Tianyuan Lu, Guillaume Butler-Laporte, Sirui Zhou, J. Brent Richards

## Abstract

**Objective:** Observational studies on cognition found that vitamin D supplementation is associated with improved cognition. Further, recent Mendelian randomization (MR) studies have shown that increased vitamin D levels, as measured by 25-hydroxyvitamin D [25(OH)D], may protect against Alzheimer’s disease. Thus, it is possible that 25OHD may protect against Alzheimer’s disease by improving cognition.

**Methods:** To evaluate this hypothesis, we began by performing an observational study by testing the association of 25OHD levels with 5 cognitive outcomes related to memory and executive function, adjusting for age, sex, BMI, smoking status and nutritional risk index in 26,787 older individuals (median age 62). Since such observational analyses can be biased by confounding, we next performed two-sample mendelian randomization (MR) analyses testing the effect of 25(OH)D on cognitive outcomes, since this method helps to protect against bias from confounding.

**Results:** Observational studies suggested a strong protective association between 25(OH)D and cognitive measures. However, in MR analyses, we found no estimated effect of 25(OH)D on these outcomes.

**Conclusion:** These findings suggest that associations between 25(OH)D levels and cognition are likely to be biased by confounding and that the relationship between 25(OH)D levels and Alzheimer’s disease is not due to the effect of 25(OH)D on cognition.

## INTRODUCTION

According to the Canadian census bureau, the number of 85 years old and older will triple over the next 25 years to 2.5 million^1^. The rate of dementia-related diseases increases exponentially for individuals 90 years and older^2^. Therefore, identifying therapeutic targets and enhancing our understanding of age-related neurodegeneration is necessary.

Some studies have demonstrated an increased prevalence of low 25-hydroxyvitamin D [25(OH)D] levels in the elderly population^3,4^. Further, cognitive decline and loss of autonomy in the elderly, which are key drivers of Alzheimer’s disease (AD) have been observed in individuals with hypovitaminosis D^5^. There is a wide range of pathways through which vitamin D may influence brain health, including inflammation, thrombosis, and growth factors^6,7^. Several observational studies have suggested a role of vitamin D in modulating neuronal development, increasing senescence, reducing sensory and social acuity^8,9,10^. Meta-analyses of the association of vitamin D and stroke incidence and dementia risk found a dose-response relationship between vitamin D and these complications^11,12^. Since vitamin D levels can be increased with relative safety using available supplements, increasing vitamin D levels could potentially help to protect against cognitive decline.

The preclinical Alzheimer’s cognitive composite cognition score^13^ (PACC) tests multiple brain function parameters. The meta-analysis from which the score was derived demonstrated a strong association of the score with the gene encoding for the apolipoprotein E ε4 polymorphism^14^. These genetic variants are the most significant risk factors for Alzheimer’s disease, which suggests that the PACC score is a helpful score to measure Alzheimer’s disease-related cognitive impairment, since it shares biological determinants with Alzheimer’s disease.

Understanding the causal effect of vitamin D on cognitive function would therefore be important to better understand widespread findings from observational study data demonstrating an association of 25(OH)D with cognitive function. However, such studies may be biased by confounding and previous work has shown that several observed associations between vitamin D levels and disease outcomes cannot be corrected by vitamin D supplementation in large-scale randomized trials^15,16,17,18,19,20,21,22^.. Mendelian randomization (MR) is a genetic epidemiology causal inference method that uses instrumental variables to reduce bias from confounding and reverse causation^23,24^. Contrasting the existing observational studies mentioned earlier, a one-sample MR study in a meta-analysis on the impact of vitamin D on cognition found no evidence for 25(OH)D levels as a causal factor for cognitive performance^25^.. Additionally, different ways to measure cognition (including memory and global cognition scores) amongst the different cohorts made the cognition measurements heterogeneous. Finally, the study employed just two SNPs (*CYP2R1, DHCR7*) associated with 25(OH)D, thus reducing the variance explained in 25(OH)D levels and, consequently statistical power. On the other hand, recent MR studies have demonstrated that increased 25(OH)D levels protect against Alzheimer’s disease^26^. Since cognition is an important risk factor for Alzheimer’s disease, it would be important to better understand if increased 25(OH)D levels were protective for cognition and if these results depended upon age.

Therefore, to better understand the relationship between 25(OH)D and cognition (determined using the PACC score), we undertook a large-scale observational study, testing the association of 25(OH)D with cognitive performance in 26,787 individuals in the Canadian Longitudinal Study of Aging (CLSA). Next, we used MR to better understand if these associations were influenced by confounding using genome-wide significant SNPs from the largest (N=443,734) GWAS of 25OHD levels^27^ as genetic instruments. We then queried their effect on the PACC score in 17,166 samples from the Canadian longitudinal study on aging (CLSA). The results provide insights into the effect of 25(OH)D levels of cognition in humans.

## METHODS

### Observational associations

Using linear regression, we tested the association of log transformed 25(OH)D with each individual cognition score (AFT, AFT2, MAT, REYI, REYII, COLTIME) and with the composite PACC score. Since 25(OH)D is confounded by various factors^28^ (anthropometric variables, lifestyle habits, and more), we identified covariates available in the CLSA cohort to add to our regression model to partially control for this potential bias. Cognition scores were adjusted for age, sex, smoking status (answered on a questionnaire with possible answers: Yes (I currently smoke)), No (I don’t smoke and I never have), Former (I don’t smoke now but I have in the past)), high nutritional risk indicator (answered on a questionnaire with possible answers: Not at high nutritional risk, and High nutritional risk), and BMI classification for adults aged 18 and over (the classification includes underweight, normal weight, overweight, obese – Class I, obese – Class II, obese – Class III). Cognition scores were inverse normal transformed. All measures were at baseline and included at most 25,357 individuals. To control for multiple testing, we considered Bonferroni significant those associations with *P < 7.1 × 10*^*-3*^ (0.05/7 (where 7 is the number of cognitive scores tested)).

### Instrumental variables for 25(OH)D

Instrumental variables were defined as independent genome-wide significant SNPs (*P < 6.6 × 10*^*-9*^) for 25(OH)D levels in 443,734 individuals. Identification of independent loci excluded high collinearity (R^2^ > 0.9) SNPs within a 20,000 kilobase window. Details on the conditionally independent approach, imputation quality control, missingness rates excluded individuals and software tools employed in the GWAS are provided in great detail in our original paper^27^. The approach yielded 138 independent SNPs. In order to ensure that we are able to recover all the variants from the outcome GWAS, we restricted our study to SNPs with a MAF > 5%. When a SNP was not present in the results for the GWAS of the PACC, AFT, COLTIME, REYI, REYI, and MAT score, we alternatively looked up a proxy SNP in linkage disequilibrium, with an R^2^ ≥ 0.8 using the 1000G European reference panel^29^ (**Supplementary Table2**). Therefore, we limited our analysis to 80 SNPs for which we estimated the variance explained in 25(OH)D. We computed the variance explained using the following formula : r^2^ = 2*β* ^2^ *ƒ* (1 –*ƒ*), where *β* and *ƒ* denote the effect of the SNP on 25OHD level and the MAF, respectively. Which we then used to calculate the *F-statistic (formula F: r*^*2*^ *x (N-2)/(1-r*^*2*^ ) where *r*^*2*^ is the variance explained and N denotes the sample size.

### Study outcomes

The PACC score is a composite score which synthesizes tests on episodic memory, timed executive function, and global cognition. The composite measure can precisely recognize early signs of cognitive changes prior to mild cognitive impairment (MCI)^13^. In terms of the definition of the individual scores, The Rey auditory verbal learning test – trial 1 and five-minute delayed recall (respectively) REYI, and REYII score represents the number of words correctly recalled in 90 seconds. The first one assesses verbal episodic memory, whereas the second one assesses delayed recall. The mental alteration test (MAT) score assesses the number of correct consecutive numeric and alphabetical alternations in 30 seconds. The time to say the ink colour of each colour name (COLTIME measured in seconds) ascertains the executive and the set switching cognitive domains. Finally, animal fluency (AFT & AFT2) assesses the different animals recited in 60 seconds. **Table 1** summarizes the individual cognitive scores in CLSA.

**Table 1.** CLSA cognitive measurements used in this study.

| **Cognitive Function** | **Cognitive Domain** | **CLSA Variable** | **Description** |
| --- | --- | --- | --- |
| Memory | Verbal Episodic Memory | The Rey Auditory Verbal Learning Test I (**REYI**) | Number of words (or variants) correctly recalled in 90 seconds – Immediate Recall |
| Memory | Delayed Recall | The Rey Auditory Verbal Learning Test II (**REYII**) | Number of words (or variants) correctly recalled in 90 seconds – Delayed Recall |
| Executive Function | Executive and Set Switching | Mental Alteration Test (**MAT**) | Number of correct consecutive numeric and alphabetical alternations in 30 seconds |
| Executive Function | Executive and Set Switching | Stroop (Victoria version) (**COLTIME**) | Time to say ink colour of each colour name (in seconds) |
| Executive Function | Verbal Fluency | Animal Fluency (**AFT1**) | Number of different animals recited in 60 seconds |
| Executive Function | Verbal Fluency | Animal Fluency (**AFT2**) | Number of different animals recited in 60 seconds |

To ascertain the association between 25(OH)D levels and the PACC^13^ score, we retrieved the effects of these 80 variants on the PACC, REYI, REYII, MAT, AFT and AFT2 scores, which is a GWAS of the aforementioned score in the CLSA cohort described elsewhere^30^. Since 92.6% of CLSA participants are of Europeans ancestry^30^, our analysis focused on individuals with European ancestry only with PACC score data (N=18,706). Population structure was identified through principal component analysis as detailed elsewhere^30^. We employed FastGWA as implemented in GCTA^31^ and PLINK2^32^ to perform association tests, which we adjusted for sex, age, and the first 20 principal components. We then stratified the population by age testing the entire cohort and individuals at least 60 years of age (N=9,489) to compare the effect of age on the cognitive scores (**Supplementary Table1**).

### Mendelian randomization

#### Units

The causal associations were evaluated using odds ratios (ORs) for the susceptibility to multiple sclerosis, which are expressed according to a standard deviation (SD) increase in 1SD of genetically predicted natural log of 25(OH)D (nmol/l). For the cognition scores, one standard deviation increase in cognitive scores are expressed according to one standard deviation increase in genetically predicted natural log of 25(OH)D (nmol/l).

#### Two-sample MR

We performed the two-sample mendelian randomization analyses using multiplicative random effect inverse-variance weighted (referred to as IVW(re)) to estimate the effect of a 1 SD increase in standardized natural log-transformed 25(OH)D on 1 a SD of PACC, AFT, REYI, REYII, MAT, COLTIME scores and odds of multiple sclerosis. We additionally stratified our analysis by age (individuals above 60 years of age versus all individuals) when estimating the effect on cognitive scores. Our analyses we performed with the package TwoSampleMR (v 0.5.6)^33^ in R(v4.2.0). We considered significant MR results with *P-value < 6.3 × 10*^*-3*^ (Bonferroni threshold accounting for multiple testing for the seven cognition scores and the risk of multiple sclerosis: 0.05/8).

#### MR Assumptions

Three assumptions are required in MR analyses. The first MR assumption requires that the genetic instrument must be strongly associated with the exposure. We selected conditionally independent genome-wide significant common (*P-value* < 6.6 × 10^−9^) variants to satisfy this assumption. Of 138 conditionally independent variants, 80 remained (**Supplementary Table1**).

The second MR assumption stipulates that genetic instruments should not be correlated to confounders linking exposure and outcome^34^.

The third MR assumption requires the instruments to influence the outcome only via their effects on the exposure (the exclusion restriction assumption), i.e. that horizontal pleiotropy does not influence the results. We therefore investigated for the presence of directional horizontal pleiotropy in our study. First, we calculated MR Egger regression slope and intercept, in addition to IVW median and IVW mode. MR-Egger is a weighted linear regression which allows the estimation of an intercept as a measure of the average pleiotropic effect and produces a slope coefficient that is robust to directional horizontal pleiotropy. MR-Egger relaxes the exclusion restriction assumption and is valid under the Instrument Strength Independent of Direct Effect (InSIDE) assumption wherein associations of the genetic variants with the 25(OH)D are independent of direct effects of the genetic variants on the cognitive score. Additionally, we performed a weighted median analysis which weights MR estimates by the effect size/standard error. This approach leverages the fact that causal estimates from non-pleiotropic instrumental variable are more probable to converge toward the median. This approach provides reliable causal estimates when less than 50% of the total weight is derived from variants with pleiotropic effects. Additionally, we assessed Cochran’s Q test. The test yields an *I2* index which gives an estimate of the amount of heterogeneity amongst IVs (*I*^*2*^ index ≤ 25%, is considered low heterogeneity, and a *I*^*2*^ index > 50% is considered high heterogeneity). Finally, we employed MR-PRESSO global test, which can detect the presence of pleiotropic outlier SNPs. By undertaking this variety of sensitivity analyses, and comparing results using these approaches, each one with different underlying assumptions, we could assess, to some degree, whether our findings were biased by horizontal pleiotropy. We considered results as significant when *P* was < 0.05 for all horizontal pleiotropy tests (MR-Egger intercept test, Cochran’s Q test, MR-PRESSO global test).

This study was conducted in accordance with the STROBE-MR guideline^29^ (STROBE-MR checklist is provided in **Supplementary Material**.

## RESULTS

### Observational associations

All the associations (**Table 2**) between log transformed 25(OH)D and the seven cognitive scores (the individual scores and the composite score) adjusted for age, sex, smoking status, nutritional index, and BMI classification were Bonferonni significant (*P-value < 7.1 × 10*^*-3*^). While the direction of effect between 25(OH)D and the cognition scores was positive, we observed a negative effect between the vitamin and the executive and set switching tests measuring how much time it takes to name ink colours (COLTIME). Thus increased 25(OH)D was consistently associated with improved cognition in observational associations.

**Table 2.** MR estimates of log(25(OH)D) on cognitive scores in CLSA. Abbreviations: IVW(re): inverse variance weighted multiplicative random effect. Wmedian: weighted median.

| Outcome | Beta (95% CI) | P-value | Outcome | Beta (95% CI) | P-value |
| --- | --- | --- | --- | --- | --- |
| PACC - Age > 60 | 9.3 x 10<sup>-4</sup> (-0.07-0.072) IVW(re) | 0.98 | PACC - All age | -0.005 (-0.058-0.048) ) IVW(re) | 0.85 |
|  | 9.6 x 10<sup>-4</sup> (-0.097-0.099) Egger | 0.98 |  | 9.6 x 10<sup>-4</sup> (-0.097-0.099) Egger | 0.87 |
|  | 0.044 (-0.056-0.14) Wmedian | 0.39 |  | 0.0038 (-0.065-0.072) Wmedian | 0.91 |
| AFT - Age > 60 | 0.13 (0.02-0.25) IVW(re) | 0.022 | AFT - All age | 0.083 (-0.0057-0.17) ) IVW(re) | 0.067 |
|  | 0.12 (-0.04-0.28) Egger | 0.15 |  | 9.6 x 10<sup>-4</sup> (-0.097-0.099) Egger | 0.11 |
|  | 0.13 (-0.024-0.29) Wmedian | 0.097 |  | 0.069 (-0.046-0.18) Wmedian | 0.24 |
| AFT2 - Age > 60 | 0.12 (0.00061-0.23) IVW(re) | 0.049 | AFT2 - All age | 0.069 (-0.022-0.16) ) IVW(re) | 0.14 |
|  | 0.088 (-0.071-0.25) Egger | 0.28 |  | 9.6 x 10<sup>-4</sup> (-0.097-0.099) Egger | 0.25 |
|  | 0.1 (-0.054-0.26) Wmedian | 0.2 |  | 0.058 (-0.06-0.17) Wmedian | 0.34 |
| COLTIME - Age > 60 | -0.047 (-0.16-0.063) IVW(re) | 0.4 | COLTIME - All age | -0.041 (-0.13-0.046) ) IVW(re) | 0.36 |
|  | 9.6 x 10<sup>-4</sup> (-0.097-0.099) Egger | 0.12 |  | 9.6 x 10<sup>-4</sup> (-0.097-0.099) Egger | 0.096 |
|  | -0.17 (-0.33--0.011) Wmedian | 0.036 |  | -0.063 (-0.18-0.055) Wmedian | 0.29 |
| MAT - Age > 60 | 0.0046 (-0.1-0.11) IVW(re) | 0.93 | MAT - All age | 0.013 (-0.064-0.09) ) IVW(re) | 0.75 |
|  | 9.6 x 10<sup>-4</sup> (-0.097-0.099) Egger | 0.91 |  | 9.6 x 10<sup>-4</sup> (-0.097-0.099) Egger | 0.76 |
|  | 0.029 (-0.13-0.19) Wmedian | 0.71 |  | -0.0085 (-0.13-0.11) Wmedian | 0.89 |
| REYI - Age > 60 | 0.021 (-0.11-0.15) IVW(re) | 0.75 | REYI - All age | 0.018 (-0.071-0.11) ) IVW(re) | 0.69 |
|  | 9.6 x 10<sup>-4</sup> (-0.097-0.099) Egger | 0.7 |  | 9.6 x 10<sup>-4</sup> (-0.097-0.099) Egger | 0.51 |
|  | 0.022 (-0.13-0.17) Wmedian | 0.78 |  | 0.015 (-0.098-0.13) Wmedian | 0.8 |
| REYII - Age > 60 | -0.02 (-0.14-0.096) IVW(re) | 0.74 | REYII - All age | 0.0076 (-0.076-0.092) ) IVW(re) | 0.86 |
|  | 0.028 (-0.13-0.19) Egger | 0.73 |  | 0.056 (-0.059-0.17) Egger | 0.34 |
|  | 0.12 (-0.044-0.29) Wmedian | 0.15 |  | 0.058 (-0.06-0.18) Wmedian | 0.34 |

### Instrumental variables for 25(OH)D

We used 80 common (MAF > 5%) independent SNPs (**S1 Table**) explaining 3.8 % of the variance in 25(OH)D levels, with an *F-statistic* of 214.75 as instruments in our MR study. 5 SNPs were absent from the PACC score GWAS and were replaced by proxies in high LD (r^2^ > 0.8). TwoSampleMR identified rs10832289, rs11182550, and rs13336347 as palindromic and were therefore removed from further analyses.

### GWAS of PACC score

We performed a genome-wide study of the PACC score in European individuals from the CLSA (N=17,166). When we restricted our analysis to individuals aged 60 and older, we found a genome-wide significant association rs429358 (*P=*1.8 × 10^−8^) at apolipoprotein E (*APOE*) (**Figure 3 C&D**). This is a well-characterized locus for dementia and Alzheimer’s disease^35,36,37^.

### PACC score and REYI, REYI, AFT, AFT2, MAT, COLTIME score outcome

For the PACC score, MR showed that the genetically predicted increase per 1 SD in natural log of 25(OH)D had no effect on PACC score estimated in the general population (Beta_PACC - All_ = -0.005, 95% CI -0.058 – 0.048, *P* = 0.85. Results were consistent in individuals over 60 (Beta_PACC – Age >60_ = 0.0009, 95% CI -0.07 – 0.07, *P* = 0.98) (**Figure 1 & Table2**).

**Figure 1.**
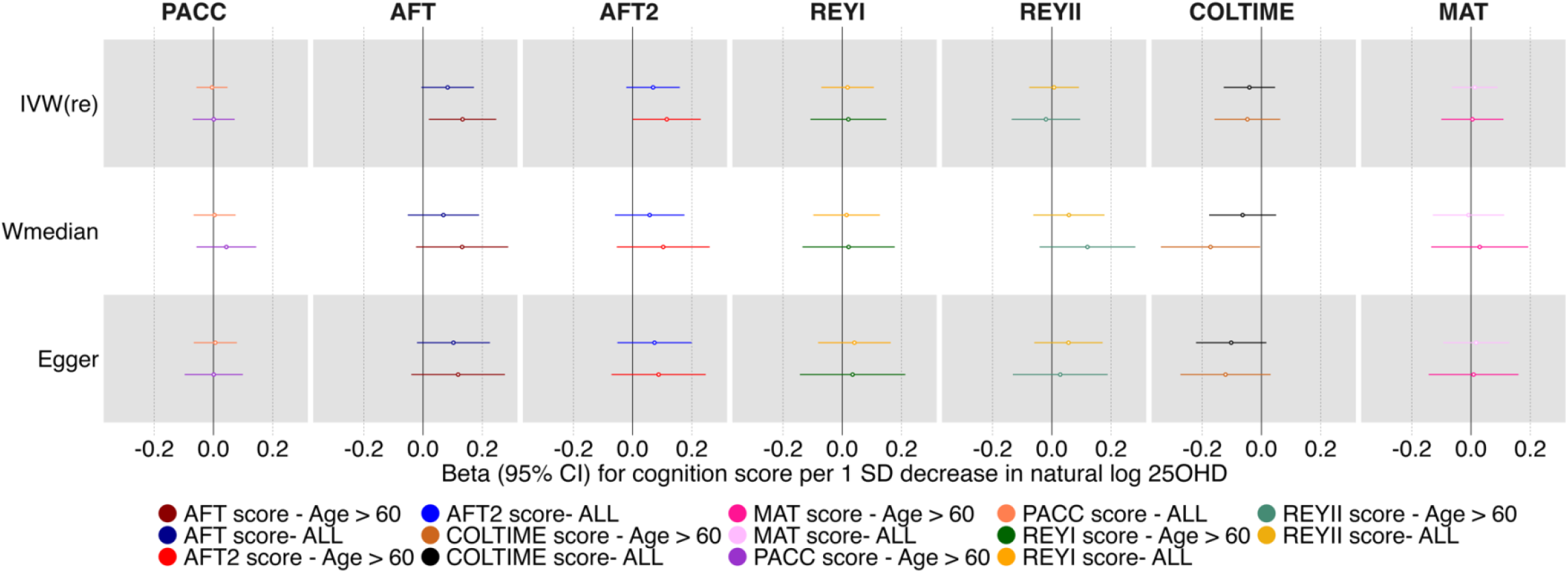
Forest plot of MR studies investigating the effect of the natural log transformed 25(OH)D on PACC, AFT, AFT2, REYI, REYII, COLTIME, and MAT scores. Forest plots of the main study and the sensitivity analyses. Significance threshold as a Bonferroni *P < 7.1 × 10*^*-3*^ Abbreviations: MR: mendelian randomization; IVW(re) random effect inverse weighted variance; Wmedian, weighted median. 25OHD, 25-hydroxyvitamin. PACC, AFT, AFT2, REYI, REYII, COLTIME, and MAT are defined in **Table1**.

**Figure 2.**
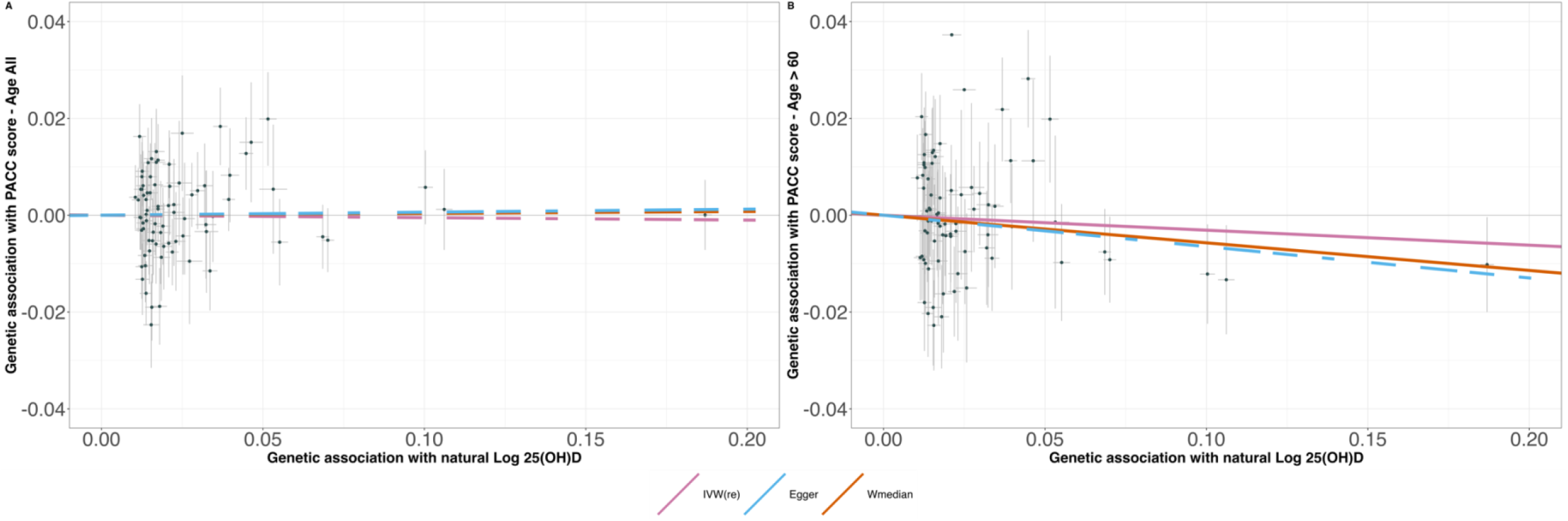
Scatter plots of the natural log 25 (OH)D MR analyses for (A) PACC score for all individuals, (B) PACC score for aged 60 years. Each dot represents a genetic instrumental variable. Three lines represent causal estimate (βIV) by the inverse variance weighted (IVW) method (pink), MR–Egger method (blue), and Weighted median method (orange). Error bars represent 95%CIs. MR, Mendelian randomization. For visualization purposes, (B) the IVW line was dashed since it overlaps with the MR-Egger line. A genetically predicted change in the natural log of 25(OH) D does not have an effect on a 1 SD of PACC score (for all individuals: beta = -0.0050, 95% CI: -0.056–0.048, *P* = 0.85; For individuals 60 years and more: beta = 0.00093, 95% CI: -0.070– 0.071, *P* = 0.98)

**Figure 3.**
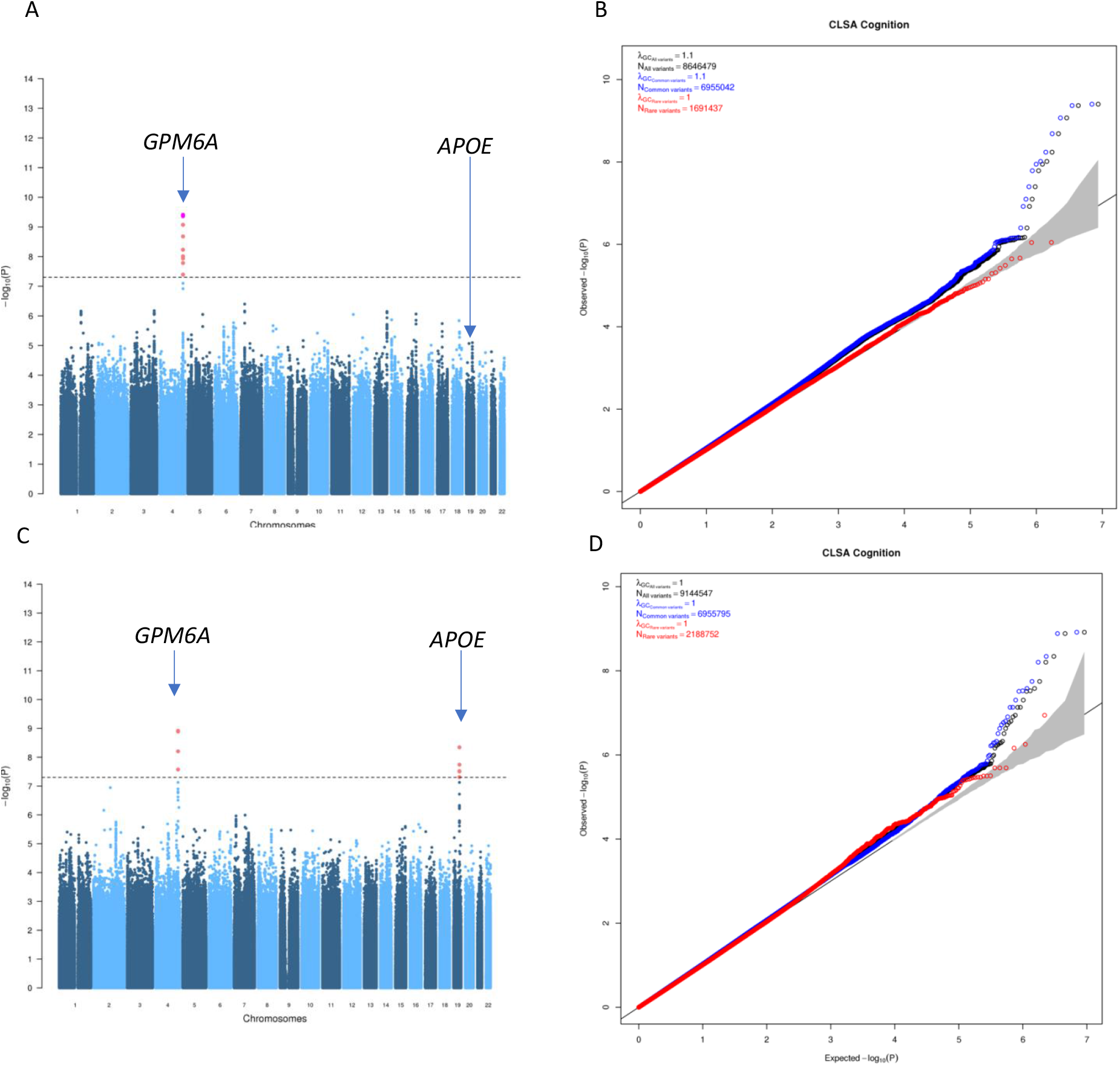
PACC score GWAS. A) Manhattan plot for PACC score (*P*_*significance*_ = 5 × 10^−8^ ; N=17,166). B) QQplot of PACC score stratified by allele frequency. C) Manhattan plot for PACC score for individuals age >= 60 (*P*_*significance*_ = 5 × 10^−8^ ; N=9,489).D) QQplot of PACC score stratified by allele frequency for individuals age >= 60. Black points are for all SNP regardless of allelic frequency, blue points are for SNPs with MAF > 1%, and red points are for SNPs with MAF < 1%. All associations are adjusted the PACC cognition score for age, sex, and the first 20 principal components.

For REYI, REYII, MAT, and COLTIME, the MR results were consistent with those from the PACC score. There was no discernible effect of genetically predicted increase per 1 SD in the natural log of 25(OH)D on any of these scores (**Table 2**).

For MR for the AFT, and AFT2 outcomes, we did observe a nominally significant association, which did not survive Bonferroni correction, in the population over 60 years of age. For AFT, the genetically predicted increase per 1 SD in natural log of 25(OH)D has no observed effect on the overall population (Beta_AFT - All_ = 0.083, 95% CI -0.0057 – 0.17, *P* = 0.07), but did show an effect in individuals over 60 (Beta_AFT2 – Age >60_ = 0.13, 95% CI 0.02 – 0.25, *P* = 0.02). The trend was consistent for AFT2, Beta_AFT2 - All_ = 0.069, 95% CI -0.022 – 0.16, *P* = 0.14, but there was an uncertain effect in individuals over 60, Beta_AFT2 – Age >60_ = 0.12, 95% CI 0.00061 – 0.23, *P* = 0.049.

### Sensitivity analysis

We computed MR-PRESSO, Egger’s regression, and Cochran’s Q test to assess the robustness of our results. Our sensitivity analyses compiled in **Table 3** for the PACC score and the other cognitive scores show that, by and large, there is low to moderate heterogeneity since *I*^2^ values are below 25%. The only moderate heterogeneity value (*I*^2^ = 47%; *I*^2^ _*Pvalue*_ = 0.005) was for the REYI score in individuals above 60 years. In fact, the MR-PRESSO global test, which can detect horizontal pleiotropic outlier SNPs, found a pleiotropic SNP for the Rey verbal learning test I. Assessing at the distortion of the causal estimate before and after outlier correction, the MR results remained the same. We found that MR-Egger intercept test, Cochran’s Q test, MR-PRESSO global test and distortion test did not detect horizontal pleiotropy (P-values > 0.05). We therefore did not identify evidence of directional horizontal pleiotropy in the MR-results.

**Table 3.**
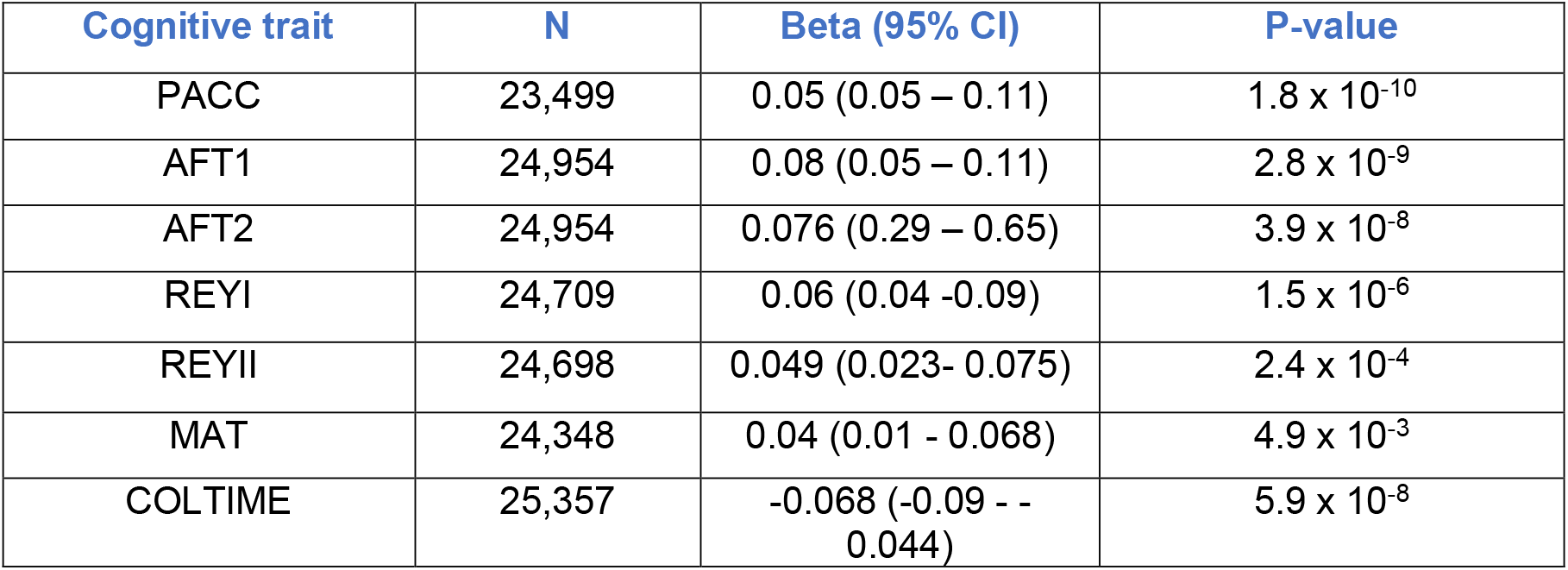
Observational associations between log(25(OH)D) and cognitive scores in CLSA. Associations were adjusted for age, sex, smoking status, BMI classification, and high nutritional risk indicator using linear regression. Scores are in 1 standard deviation, and vitamin D is Log 25(OH)D. Abbreviations: AFT: Number of different animals recited in 60 seconds; REYI: Number of words (or variants) correctly recalled in 90seconds (immediate recall); REYII: Number of words recalled in 90 seconds (delayed recall); MAT: Number of correct consecutive numeric and alphabetical alternations in 30 seconds; COLTIME: Time to say ink color of each color name (in seconds); N: sample size; Beta (95%CI): effect size of Log 25(OH)D on standard deviation of each cognitive score. 95% confidence intervals are displayed in paratheses.

**Table 4.** MR sensitivity analysis.

| Exposure | Outcome | Cochran Q |  | MR-Egger |  | MRPRESSO |  |  |  |  |
| --- | --- | --- | --- | --- | --- | --- | --- | --- | --- | --- |
|  |  | I<sup>2</sup> | I<sup>2</sup> p-value | Egger intercept | P-value Egger intercept | Causal Estimate | P-value | Outlier detected | Outlier corrected estimate | Outlier corrected p-value |
| 25(OH)D | PACC - Age > 60 | 19.7 | 0.12 | -1.80 x 10<sup>-6</sup> | 1 | 0.00093 | 0.98 | 0 | 0 | 0 |
| 25(OH)D | PACC- All age | 19.7 | 0.11 | -6.00 x 10<sup>-4</sup> | 0.66 | 0.00016 | 0.99 | 0 | 0 | 0 |
| 25(OH)D | AFT2_score - Age > 60 | 13.2 | 0.21 | 1.5 x 10<sup>-3</sup> | 0.62 | 0.12 | 0.052 | 0 | 0 | 0 |
| 25(OH)D | AFT2_score - All age | 27.6 | 0.056 | -2.6 x 10<sup>-4</sup> | 0.91 | 0.069 | 0.13 | 0 | 0 | 0 |
| 25(OH)D | AFT_score - Age > 60 | 10.5 | 0.24 | 7.9 x 10<sup>-4</sup> | 0.79 | 0.13 | 0.024 | 0 | 0 | 0 |
| 25(OH)D | AFT_score- All age | 22.3 | 0.087 | -1.0 x 10<sup>-3</sup> | 0.65 | 0.08 | 0.07 | 0 | 0 | 0 |
| 25(OH)D | COLTIME_score - Age > 60 | 10.5 | 0.25 | 3.9 x 10<sup>-3</sup> | 0.17 | -0.048 | 0.4 | 0 | 0 | 0 |
| 25(OH)D | COLTIME_score- All age | 21.1 | 0.097 | 3.3 x 10<sup>-3</sup> | 0.14 | -0.04 | 0.36 | 0 | 0 | 0 |
| 25(OH)D | MAT_score - Age > 60 | 0 | 0.69 | -2.2 x 10<sup>-4</sup> | 0.94 | 0.0046 | 0.93 | 0 | 0 | 0 |
| 25(OH)D | MAT_score- All age | 0 | 0.71 | -2.5 x 10<sup>-4</sup> | 0.9 | 0.01 | 0.74 | 0 | 0 | 0 |
| 25(OH)D | REYI_score - Age > 60 | 47.4 | 0.0049 | -7.5 x 10<sup>-4</sup> | 0.82 | 0.021 | 0.75 | 1 | 0.031 | 0.61 |
| 25(OH)D | REYI_score- All age | 26.3 | 0.057 | -1.2 x 10<sup>-3</sup> | 0.59 | 0.018 | 0.69 | 0 | 0 | 0 |
| 25(OH)D | REYII_score - Age > 60 | 22.4 | 0.095 | -2.6x 10<sup>-3</sup> | 0.39 | -0.02 | 0.74 | 0 | 0 | 0 |
| 25(OH)D | REYII_score- All age | 15.8 | 0.16 | -2.6x 10<sup>-3</sup> | 0.23 | 0.0076 | 0.86 | 0 | 0 | 0 |

## DISCUSSION

In this study, we carried out an observational study followed by a two-sample MR to ascertain the effect of 25(OH)D supplementation on cognitive outcome test scores. First, we computed the linear regression adjusting for age, sex, BMI, smoking status, and nutritional risk index, between 25(OH)D and cognition test scores. Higher 25(OH)D levels were associated with improvements in all cognition measures. We then used MR, which reduces bias from reverse causation and confounding, thus enabling us to test for causal effects. The results of the MR study did not support a causal effect between a genetically determined change in 25(OH)D levels and the PACC score and the cognitive score. These results suggest that observational associations between 25(OH)D levels and improved cognition are likely due to confounding.

Vitamin D supplementation has been proposed for treatment of many common diseases. While supplementation could be beneficial in certain contexts^15^ (i.e. prevention of multiple sclerosis, hypovitaminosis D and Alzheimer’s disease), results from observational studies need to be considered carefully. For instance, an observational study published in 2014^38^ with a diverse and small cohort (N=382; 41.4% individuals from European ancestry, 29.6% individuals from African American ancestry, 25.1% individuals from Latino/Hispanic ancestry) showed that low vitamin D level is associated with an increased rate of cognitive decline. However, this was not supported by an MR study in a cohort of 153,187 individuals of European ancestry. The study looked at the effect of only two 25(OH)D canonical variants rs12785878 (vitamin D-decreasing allele, G), located near gene coding 7-dehydrocholesterol reductase (DHCR7), and rs12794714 (vitamin D-decreasing allele, A) near 25-hydroxylase (CYP2R1) on global cognition score (Beta: - 0.00; 95% CI: (−0.01, 0.01); P-value = 1.0), and memory cognitive score (Beta: -0.00; 95% CI: (−0.01, 0.00); P-value = 0.6). Both the previously published observational studies and MR studies on the effect of 25(OH)D on cognitive scores are congruent with our study^25^, ^38^.

How 25(OH)D may protect against Alzheimer’s disease in human is unknown. MR has shown a protective effect of 25(OH)D on Alzheimer’s disease in humans^26,39,40^. Additionally, previous and novel evidence demonstrated that cognition is protective for Alzheimer’s disease^40^, ^41^. Therefore, we hypothesized that 25(OH)D may protect against Alzheimer’s disease in humans by increasing cognition. However, our results suggest that the observed MR effects of 25(OH)D on Alzheimer’s disease are explained by improved cognition.

We note several strengths in our study. We leveraged the strengths of the MR design to lessen bias from confounders and reverse causation, therefore enabling more reliable estimate causal effects provided adherence to MR assumptions. To maximize statistical power, we utilized cohorts with large sample sizes.

Our study also contains several limitations. Since MR depends mainly on three causal inference assumptions (relevance, independence, and exclusion restriction) any violation of these could invalidate our results. To address these assumptions, we performed several sensitivity analyses investigating horizontal pleiotropy and heterogeneity (MR-Egger, MR-PRESSO, Cochran’s Q test). Our causal estimate between 25(OH)D and REYI score in older individuals was considered to have a moderate amount of heterogeneity. After the removal of outlying SNP through MR-PRESSO, the causal estimate remained the same. This suggests that the MR results are not likely to be strongly biased by directional horizontal pleiotropy. Further, our study focuses on European ancestry individuals, therefore, conclusions and interpretation of the results are not generalizable to other ancestries. This once again emphasizes the need for large-scale studies assessing the genetic determinants of disease in individuals of non-European ancestry.

In conclusion, our MR study found no evidence for a causal effect between 25(OH)D concentrations and cognitive performance in European ancestry individuals.

## Supporting information

Stables_1_2

## Data Availability

All data produced are available online at the GWAS catalog

http://ftp.ebi.ac.uk/pub/databases/gwas/summary_statistics/GCST90239001-GCST90240000//GCST90239732/

http://ftp.ebi.ac.uk/pub/databases/gwas/summary_statistics/GCST90239001-GCST90240000//GCST90239733/

http://ftp.ebi.ac.uk/pub/databases/gwas/summary_statistics/GCST010001-GCST011000/GCST010144

## FUNDING STATEMENT

The Richards research group is supported by the Canadian Institutes of Health Research (CIHR: 365825; 409511, 100558, 169303), the McGill Interdisciplinary Initiative in Infection and Immunity (MI4), the Lady Davis Institute of the Jewish General Hospital, the Jewish General Hospital Foundation, the Canadian Foundation for Innovation, the NIH Foundation, Cancer Research UK, Genome Québec, the Public Health Agency of Canada, McGill University, Cancer Research UK [grant number C18281/A29019] and the Fonds de Recherche Québec Santé (FRQS). JBR is supported by a FRQS Mérite Clinical Research Scholarship. Support from Calcul Québec and Compute Canada is acknowledged. TwinsUK is funded by the Welcome Trust, Medical Research Council, European Union, the National Institute for Health Research (NIHR)-funded BioResource, Clinical Research Facility and Biomedical Research Centre based at Guy’s and St Thomas’ NHS Foundation Trust in partnership with King’s College London. These funding agencies had no role in the design, implementation or interpretation of this study.

## CONFLICT OF INTEREST

JBR’s institution has received investigator-initiated grant funding from Eli Lilly, GlaxoSmithKline and Biogen for projects unrelated to this research. He is the CEO of 5 Prime Sciences (www.5primesciences.com), which provides research services for biotech, pharma and venture capital companies for projects unrelated to this research.

## ACKNOWLEDGMENTS

We thank all the participants, the UK Biobank coordinating team, and the CLSA (Project# 1906026 and 190208) coordinating team for their approval, valuable time, generosity, and contributions to the data collection.

## ETHICS STATEMENT

Ethical review and approval was not required for the study on human participants in accordance with the local legislation and institutional requirements. The patients/participants provided their written informed consent to participate in this study.

## DATA AVAILABILITY

All GWAS summary statistics used in this study are made publicly available through the GWAS catalog. 25(OH)D and PACC score GWAS summary statistics (All ages and individuals 60 and older) can be downloaded through the following accession: 25(OH)D: GCST010144; PACC_GWAS: GCST90239732; PACC_GWAS_old: GCST90239733. Further inquiry can be directed to the corresponding author.

## Notes

### Author Declarations

Ethics committee/IRB of Canadian Institutes of Health Research (CIHR) Advisory Committee on Ethical, Legal, and Social Issues (ELSI) for the CLSA gave ethical approval for this work

## References

1. Statistics Canada. Census in Brief A portrait of Canada’s growing population aged 85 and older from the 2021 Census. Published online April 27, 2022. Accessed September 8, 2022. https://www12.statcan.gc.ca/census-recensement/2021/as-sa/98-200-X/2021004/98-200-X2021004-eng.pdf

2. Corrada MM, Brookmeyer R, Paganini-Hill A, Berlau D, Kawas CH. Dementia incidence continues to increase with age in the oldest old: The 90+ study. Ann Neurol. 2010;67(1):114-121. doi:https://doi.org/10.1002/ana.21915

3. Remelli F, Vitali A, Amedeo Z, Volpato S. Vitamin D Deficiency and Sarcopenia in Older Persons. Nutrients. Published online 2019. doi:https://doi.org/10.3390/nu11122861

4. Cédric A, Olivier B. Vitamin D in Older Adults: The Need to Specify Standard Values with Respect to Cognition. Published online April 15, 2014. doi:10.3389/fnagi.2014.00072

5. Thorleif E, Sander D, Horst B, Kerstin S, Förstl H. Vitamin D deficiency, cognitive impairment and dementia: a systematic review and meta-analysis. Dement Geriat Cogn Disord. Published online 2012. doi:doi: 10.1159/000339702

6. Gold J, Shoaib A, Gorthy G, Grossberg GT. The Role of Vitamin D in Cognitive Disorders in Older Adults. US Neurol. 2018;14(1):41. doi:10.17925/USN.2018.14.1.41

7. Nitsa A, Toutouza M, Machairas N, Mariolis A, Philippou A, Koutsilieris M. Vitamin D in Cardiovascular Disease. In Vivo. 2018;32(5):977-981. doi:10.21873/invivo.11338

8. Keisala T, Minasyan A, Lou YR, et al. Premature aging in vitamin D receptor mutant mice. J Steroid Biochem Mol Biol. 2009;115(3-5):91-97. doi:10.1016/j.jsbmb.2009.03.007

9. Brown J, Bianco JI, McGrath JJ, Eyles DW. 1, 25-Dihydroxyvitamin D(3) induces nerve growth factor, promotes neurite outgrowth and inhibits mitosis in embryonic rat hippocampal neurons. Neurosci Lett. Published online 2003. doi:10.1016/S0304-3940(03)00303-3

10. Goodwill AM, Szoeke C. A Systematic Review and Meta-Analysis of The Effect of Low Vitamin D on Cognition. J Am Geriatr Soc. 2017;65(10):2161-2168. doi:10.1111/jgs.15012

11. Jayedi A, Rashidy-Pour A, Shab-Bidar S. Vitamin D status and risk of dementia and Alzheimer’s disease: A meta-analysis of dose-response †. Nutr Neurosci. 2019;22(11):750-759. doi:10.1080/1028415X.2018.1436639

12. Shi H, Chen H, Zhang Y, et al. 25-Hydroxyvitamin D level, vitamin D intake, and risk of stroke: A dose-response meta-analysis. Clin Nutr Edinb Scotl. 2020;39(7):2025-2034. doi:10.1016/j.clnu.2019.08.029

13. Donohue MC, Sperling RA, Salmon DP, et al. The preclinical Alzheimer cognitive composite: Measuring amyloid-related decline. JAMA Neurol. 2014;71(8):961-970. doi:10.1001/jamaneurol.2014.803

14. Liu CC, Kanekiyo T, Xu H, Bu G. Apolipoprotein E and Alzheimer disease: risk, mechanisms and therapy. Nat Rev Neurol. 2013;9(2):106-118. doi:10.1038/nrneurol.2012.263

15. Bouillon R, Manousaki D, Rosen C, Trajanoska K, Rivadeneira F, Richards JB. The health effects of vitamin D supplementation: evidence from human studies. Nat Rev Endocrinol. 2022;18(2):96-110. doi:10.1038/s41574-021-00593-z

16. Manson JE, Cook NR, Lee IM, et al. Vitamin D Supplements and Prevention of Cancer and Cardiovascular Disease. N Engl J Med. Published online 2018:NEJMoa1809944. doi:10.1056/NEJMoa1809944

17. Manson JE, Bassuk SS, Cook NR, et al. Vitamin D, Marine n-3 Fatty Acids, and Primary Prevention of Cardiovascular Disease Current Evidence. Circ Res. 2020;126(1):112-128. doi:10.1161/CIRCRESAHA.119.314541

18. Djoussé L, Cook NR, Kim E, et al. Supplementation With Vitamin D and Omega-3 Fatty Acids and Incidence of Heart Failure Hospitalization: VITAL-Heart Failure. Circulation. 2020;141(9):784-786. doi:10.1161/CIRCULATIONAHA.119.044645

19. Chandler PD, Chen WY, Ajala ON, et al. Effect of Vitamin D 3 Supplements on Development of Advanced Cancer: A Secondary Analysis of the VITAL Randomized Clinical Trial. JAMA Netw Open. 2020;3(11):e2025850. doi:10.1001/jamanetworkopen.2020.25850

20. Manson JE, Bassuk SS, Buring JE. Principal results of the VITamin D and OmegA-3 TriaL (VITAL) and updated meta-analyses of relevant vitamin D trials. J Steroid Biochem Mol Biol. 2020;198:105522. doi:10.1016/j.jsbmb.2019.105522

21. Christen WG, Cook NR, Manson JE, et al. Effect of Vitamin D and ω-3 Fatty Acid Supplementation on Risk of Age-Related Macular Degeneration: An Ancillary Study of the VITAL Randomized Clinical Trial. JAMA Ophthalmol. 2020;138(12):1280. doi:10.1001/jamaophthalmol.2020.4409

22. Bassuk SS, Chandler PD, Buring JE, Manson JE, for the VITAL Research Group. The VITamin D and OmegA-3 TriaL (VITAL): Do Results Differ by Sex or Race/Ethnicity? Am J Lifestyle Med. 2021;15(4):372-391. doi:10.1177/1559827620972035

23. Burgess S, Butterworth A, Thompson SG. Mendelian randomization analysis with multiple genetic variants using summarized data. Genet Epidemiol. 2013;37(7):658-665. doi:10.1002/gepi.21758

24. Davey Smith G, Ebrahim S. ‘Mendelian randomization’: can genetic epidemiology contribute to understanding environmental determinants of disease?*. Int J Epidemiol. 2003;32(1):1-22. doi:10.1093/ije/dyg070

25. Maddock J, Zhou A, Cavadino A, et al. Vitamin D and cognitive function: A Mendelian randomisation study. Sci Rep. 2017;7(1):13230. doi:10.1038/s41598-017-13189-3

26. Mokry LE, Ross S, Morris JA, Manousaki D, Forgetta V, Richards JB. Genetically decreased vitamin D and risk of Alzheimer disease. Neurology. Published online 2016. doi:10.1212/WNL.0000000000003430

27. Manousaki D, Mitchell R, Dudding T, et al. Genome-wide Association Study for Vitamin D Levels Reveals 69 Independent Loci. Am J Hum Genet. 2020;106(3):327-337. doi:10.1016/j.ajhg.2020.01.017

28. Datta S, Pal M, De A. The dependency of vitamin d status on anthropometric data. Malays J Med Sci MJMS. 2014;21(3):54–61.

29. Skrivankova VW, Richmond RC, Woolf BAR, et al. Strengthening the reporting of observational studies in epidemiology using mendelian randomisation (STROBE-MR): explanation and elaboration. BMJ. Published online October 26, 2021:2233. doi:10.1136/bmj.n2233

30. Forgetta V, Li R, Darmond-Zwaig C, et al. Cohort profile: genomic data for 26 622 individuals from the Canadian Longitudinal Study on Aging (CLSA). BMJ Open. 2022;12(3):e059021. doi:10.1136/bmjopen-2021-059021

31. Yang J, Lee SH, Goddard ME, Visscher PM. GCTA: a tool for genome-wide complex trait analysis. Am J Hum Genet. 2011;88(1):76-82. doi:10.1016/j.ajhg.2010.11.011

32. Chang CC, Chow CC, Tellier LC, Vattikuti S, Purcell SM, Lee JJ. Second-generation PLINK: rising to the challenge of larger and richer datasets. GigaScience. 2015;4(1):7. doi:10.1186/s13742-015-0047-8

33. Hemani G, Zheng J, Elsworth B, et al. The MR-Base platform supports systematic causal inference across the human phenome. eLife. 2018;7:e34408. doi:10.7554/eLife.34408

34. Vimaleswaran KS, Berry DJ, Lu C, et al. Causal Relationship between Obesity and Vitamin D Status: Bi-Directional Mendelian Randomization Analysis of Multiple Cohorts. Minelli C, ed. PLoS Med. 2013;10(2):e1001383. doi:10.1371/journal.pmed.1001383

35. Lambert JC, Ibrahim-Verbaas CA, Harold D, et al. Meta-analysis of 74,046 individuals identifies 11 new susceptibility loci for Alzheimer’s disease. Nat Genet. 2013;45(12):1452-1458. doi:10.1038/ng.2802

36. Naj AC, Jun G, Beecham GW, et al. Common variants at MS4A4/MS4A6E, CD2AP, CD33 and EPHA1 are associated with late-onset Alzheimer’s disease. Nat Genet. 2011;43(5):436-441. doi:10.1038/ng.801

37. Deelen J, Evans DS, Arking DE, et al. Publisher Correction: A meta-analysis of genome-wide association studies identifies multiple longevity genes. Nat Commun. 2021;12(1):2463. doi:10.1038/s41467-021-22613-2

38. Miller JW, Harvey DJ, Beckett LA, et al. Vitamin D Status and Rates of Cognitive Decline in a Multiethnic Cohort of Older Adults. JAMA Neurol. 2015;72(11):1295. doi:10.1001/jamaneurol.2015.2115

39. Wang L, Qiao Y, Zhang H, et al. Circulating Vitamin D Levels and Alzheimer’s Disease: A Mendelian Randomization Study in the IGAP and UK Biobank. Yu JT, ed. J Alzheimers Dis. 2020;73(2):609-618. doi:10.3233/JAD-190713

40. Larsson SC, Traylor M, Malik R, Dichgans M, Burgess S, Markus HS. Modifiable pathways in Alzheimer’s disease: Mendelian randomisation analysis. BMJ. Published online December 6, 2017:j5375. doi:10.1136/bmj.j5375

41. Hu Y, Zhang Y, Zhang H, et al. Cognitive performance protects against Alzheimer’s disease independently of educational attainment and intelligence. Mol Psychiatry. Published online July 15, 2022. doi:10.1038/s41380-022-01695-4

